# Utility and limitations of Google searches for tracking disease: the case of taste and smell loss as markers for COVID-19

**DOI:** 10.1101/2020.05.07.20093955

**Authors:** Kim Asseo, Fabrizio Fierro, Yuli Slavutsky, Johannes Frasnelli, Masha Y Niv

## Abstract

Web tools are widely used among population to obtain health related information, and these data are often employed for public health monitoring. Here we analyzed searches related to smell loss and taste loss, recently linked to COVID-19, as well as sight loss and hearing loss, not included in the COVID-19 symptoms list. Google Trends results per region (Italy) or state (United States) over several weeks were compared to the number of new cases prevalence in that geographical area. Taste and smell loss searches were correlated with each other, and, during a limited time window, with new COVID-19 cases. However, this correlation decreased with time, attributable, at least in part, to media coverage. As new symptoms are being discovered for COVID-19 and the pandemic continues to spread around the globe, the lesson learned here, that correlation between public interest in novel symptoms of infectious disease has an initial spike (the “surprise rise”) and subsequently goes to a new baseline due to “knowledge saturation”, is of general and practical value for the public.

## Introduction

SARS-CoV-2 pandemic has by now hit almost all countries worldwide. Monitoring disease occurrence is a key prerequisite for combating its spread. Laboratory testing availability differs from country to country, most nations not being able to test the general and even the symptomatic population broadly. Furthermore, symptoms elicited by SARS-CoV-2 are still being discovered, with the list of officially recognized symptoms being updated on a rolling basis.

Specifically, in addition to fever, cough, shortness of breath or difficulty breathing, chill, muscle pain, headache, and sore throat, recent additions to [CDC-listed symptoms](https://www.cdc.gov/coronavirus/2019-ncov/symptoms-testing/symptoms.html) include “new taste and smell loss”. The [National Health Services](https://www.nhs.uk/conditions/coronavirus-covid-19/check-if-you-have-coronavirus-symptoms/) of the UK describes “loss or change to your sense of smell or taste” among the symptoms and the [World Health Organization](https://www.who.int/health-topics/coronavirus#tab=tab_3) has listed “loss of taste or smell” among “less common symptoms” of COVID-19. Smell loss (anosmia), and to a lesser degree taste loss (ageusia), accompanying COVID-19 infection have appeared in reports of COVID-19 patients’ testimonies^1^, preprints of scientific papers^2-4^, peer-reviewed publications (e.g.^5-9^), and are being widely discussed by journalists.

The details of taste and smell change in relation to COVID-19 remain unclear, and scientists (including the authors of this contribution), clinicians and patients’ advocates have created a dedicated consortium (<u>GCCR</u>) which studies the relation between taste and smell loss and COVID-19.

Here we set out to explore the hypothesis, proposed by several groups ^10-12^ and also discussed in The New York Times^13^ and on CNBC^14^, that Google Trends searches on smell loss are indicative of COVID-19 cases.

We focused on two countries, Italy and the US, and have looked not only for searches on smell loss, but also for those of taste loss. Media reports on these phenomena were analyzed using Media Cloud a database collecting digital media articles (hence, not including radio, television or printed media) and quantifying the attention over time to a query topic.

Furthermore, we used sight loss and hearing loss as controls, since these are senses not currently known to be associated with COVID-19. Because smell and taste loss are gradually becoming recognized as COVID-19 symptoms, we also looked at searches for COVID-19 symptoms in general.

## Results

We analyzed Google searches and the numbers of new COVID-19 cases in Italy and the US, considering the states (US) and the regions (Italy) they are composed by. For each region or state, the following parameters were calculated: Google Trends popularity index per region/state for generic (COVID-19 symptoms) and specific (taste loss, smell loss) symptoms of COVID-19, as well as specific symptoms not known to affect patients (hearing loss, sight loss); the number of digital media articles mentioning the smell loss and taste loss symptoms, normalized per the total number of digital media articles analyzed; the number of new COVID-19 cases per region or state normalized per 1,000,000 inhabitants. The data were collected for eleven consecutive weeks, spanning March 4th to May 19th, 2020. Next, for each week and for each of the searches, the correlation with the normalized number of new cases was calculated for each country.

Results for weeks with a good correlation for taste loss and smell loss searches are shown in Figure 1. The volume of searches for these two keywords was high, among the other regions, in Lombardy, Emilia Romagna and Veneto, as well as New York, New Jersey and Louisiana, which are geographical sub-areas with high rates of new COVID-19 patients/inhabitants in their respective country.

**Figure 1:**
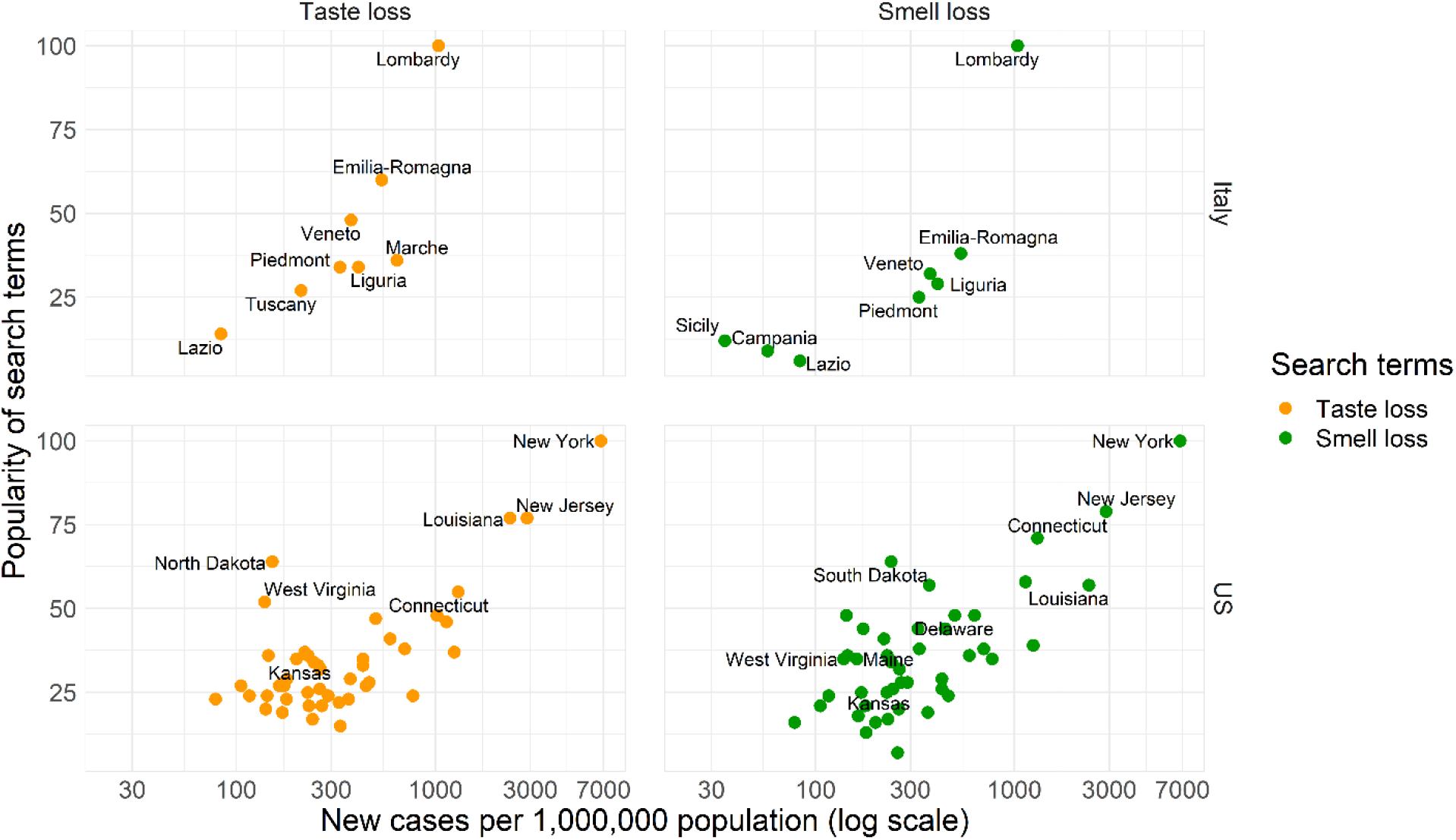
Data from which correlation was derived. Here shown for Italy (11-17 March) and the US (1-7 April), the weeks with highest correlation for taste loss. The graphs show the taste loss (in orange) and smell loss (in green) search queries. Each point represents a different region/state. Normalized number of new cases related to the corresponding week on the x axis, popularity of the search terms for the same week on the y axis. For both Italy and the US graphs, not all the regions or states are shown because of the lack of popularity index for some regions/states.

Does this picture hold over time? In Figure 2, we followed week by week the correlation between the number of searches and the number of new cases, calculated for 20 regions in Italy and 51 states in the US (as was exemplified for a particular week in Figure 1).

**Figure 2:**
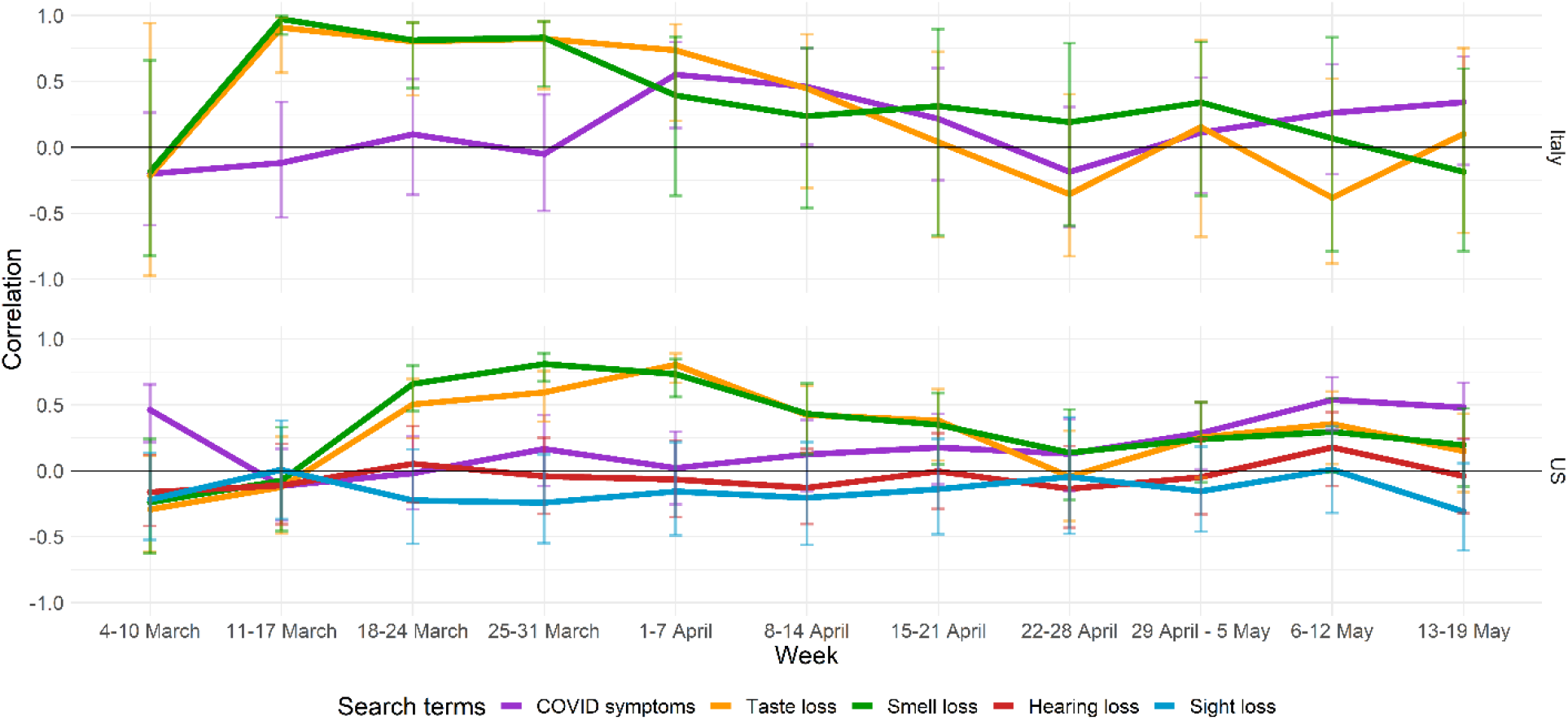
Weekly correlation data. Correlation values between the number of new COVID-19 cases, normalized with respect to 1,000,000 inhabitants, in the monitored regions of Italy (top graph) and states of the US (bottom graph), and search queries popularity index, for each week considered. Confidence intervals are shown as error bars.

Overall, Figure 2 illustrates that both in Italy and in the US, there were some weeks with a strong correlation between the normalized number of new cases and the number of searches for taste and smell loss. This may be attributable to people affected by the taste loss and/or smell loss searching for the specific symptoms they have experienced. However, this correlation changed over time: during the first analyzed week in Italy (4-10 March) and the first two in the US (4-10 and 11-17 March), taste loss and smell loss queries showed a low negative correlation with new COVID-19 cases. In the following weeks, the peaks with the highest correlation values were reached for Italy in the 11-17 March week (0.91 for taste loss and 0.97 for smell loss) and for the US in the 25-31 March and 1-7 April weeks (0.81 for both smell loss and taste loss). The weeks following the peaks are characterized by a decrease in correlation for these two symptom-specific keywords and, despite oscillations, the values do not reach the peaks again (see Supplementary Fig. S1 for a representative example of data for a week with low correlation (22-28 April)). In Italy, in the 11-17 March and 15-21 April weeks, the average number of new cases per 1,000,000 inhabitants was almost identical, 354 and 357, respectively, while the correlation dropped from 0.91 for taste loss and 0.97 for smell loss to 0.04 and 0.31, respectively (Figure 2). Similarly, from 1-7 April week to the 22-28 April week, the average number of new cases in the US remained confined within the range of 644-663, while correlation decreased from 0.81 for taste loss and 0.73 for smell loss to −0.04 and 0.14 respectively.

Hearing loss and sight loss search terms, which were used as controls, show lack of correlation in the US in all of the eleven weeks considered in the analysis, as expected (Figure 2). The corresponding searches for the Italian translation of the queries (“perdita udito” and “perdita vista”) did not produce enough results to show data relative to different regions and, consequently, display no correlation. Conjunctivitis has been recently added by the WHO to the list of the less common COVID-19 symptoms, but the low correlation with sight loss searches supports the hypothesis that it does not usually interfere with eyesight^15^. Only a low correlation in some of the eleven weeks studied here was observed for “COVID-19 symptoms” search. This can be attributed to a general interest in the disease and its symptoms, with some contribution of searches by people suspecting they might have contracted the disease. The moderate rise in correlation with “COVID-19 symptoms” searches observed in the last weeks may be due to the ongoing updates of new symptoms and current lack of understanding of this disease^16^.

Since we hypothesized that media coverage of COVID-19 related taste and smell symptoms may impact the number of searches, we next analyzed, on a daily basis and for each country as a whole, the number of new cases, as well as that of Google searches and the volume of digital media coverage of taste and smell loss.

Using the Explorer tool on the Media Cloud platform, we monitored the number of times taste and smell keywords were mentioned daily by digital news media.

The news about these two sensory-related symptoms, in both Italy and the US, was reported for the first time during the 11-17 March week, according to our search on Media Cloud (media coverage at the end of the 4-10 March week in both Italy and the US actually reported the taste loss or smell loss in a different context, not related to COVID-19 symptoms) (Figure 3). The maximum media coverage in Italy was reached between the 23rd and 24th of March, with a second higher peak for smell loss reporting on April 12th. During the 11-17 March week, the correlation peak for Italy, reported in Figures 1 and 2, was reached. An increase in popularity of searches was observed before the first media reports of March 14th (Figure 3 and Supplementary Fig. S2). Indeed, the total number of cases on March 13th in Italy was 8.2 times higher than in the US (17,660 for Italy vs 2,147 for the US) and, consequently, the volume of searches observed up to that date was also higher in Italy (Supplementary Fig. S2).

**Figure 3:**
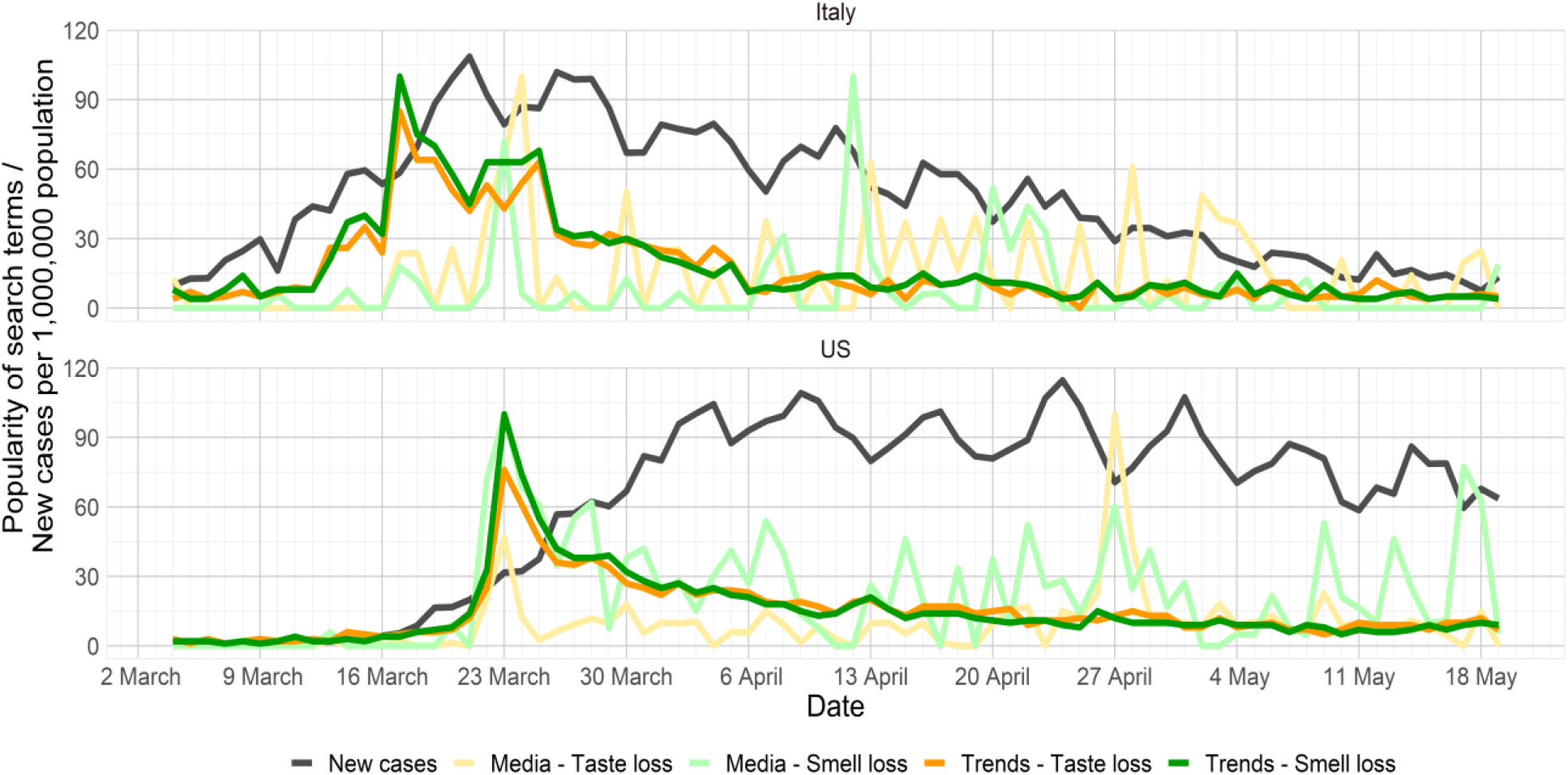
Media coverage, the number of new cases and Google searches over time. Comparison between Google searches volume for taste loss and smell loss queries, Media Cloud popularity of the same keywords and the number of new COVID-19 cases. Calculation was performed from March 4th to May 19th for Italy and the US. The Media Cloud data were normalized as for the Google search results, assigning the value of 100 to the day with the highest popularity, with the other days assigned values relative to that day. The number of new cases is relative to a population of 1,000,000 in the respective country.

The scenario we uncovered for the US was different. The Media Cloud data and the Google Trends searches are perfectly superimposed in proximity of the first high peaks. The rise of cases in the US set in later than in Italy, and there was no time window in which the number of cases was surging while the taste and smell symptoms were still unknown. Therefore, in the case of the US, it is difficult to know which part of taste and smell loss searches were driven by patients worried about their own symptoms, and which part was due to interest elicited by the media.

A correlation between the number of new cases and the volume of searches is more pronounced in Italy than in the US, while the volume of searches in the US is more affected by the media trends than in Italy (Table 1). Smell and taste loss trends are more closely related to each other than to the respective media coverage of the individual effect, suggesting that the two symptoms are experienced in concert. This is of interest, as it may indirectly suggest a potential common or joint mechanism that allows SARS-CoV-2 to affect both senses together.

**Table 1:** Pair-wise correlation. Calculated pair-wise correlation between the variables (left and right in the correlation variables column) in Italy and the US, as visualized in Figure 3.

| Correlation variables |  | Italy | US |
| --- | --- | --- | --- |
| **Trends - Taste loss** | **Trends - Smell loss** | **0.93** | **0.97** |
| Media - Taste loss | Trends - Taste loss | 0.21 | 0.74 |
| Media - Smell loss | Trends - Smell loss | 0.04 | 0.79 |
| New cases | Trends - Taste loss | 0.61 | 0.27 |
| New cases | Trends - Smell loss | 0.59 | 0.19 |

Overall, the decrease in correlation between taste and smell loss searches and the number of new cases over time is apparent both for Italy and the US (Figure 2). We believe that, with media coverage and inclusion of taste and smell loss as official symptoms of COVID-19, less searches are being conducted for these symptoms taken in isolation. Rather, a convergence with a set of searches for the established COVID-19 symptoms is observed, which is attributable to a better knowledge of the pertinent symptoms by the general population.

## Discussion

In this study, we showed the existence of a correlation between Google searches for specific new symptoms of COVID-19 (taste loss and smell loss) and the number of people affected by the SARS-CoV-2 virus in Italy and the US. In general, regions/states with a high percentage of infection cases tend to search for these specific symptoms more often than those with a low incidence when the number of new cases reach a relatively high volume (21,357 for Italy on 11-17 March and 47,553 for the US on 18-24 March) for the first time. Nevertheless, relying solely on Google trends for taste loss and smell loss searches to monitor the spread of the SARS-CoV-2, as suggested by Goldman and Sachs^14^, is not a reliable strategy. Indeed, our analysis showed that this correlation varies from week to week, even when the incidence of new cases in a country is high. Notably, the correlation weakens with time, the most recent weeks showing a vanishing correlation - possibly because the specific gustatory and olfactory symptoms are becoming more broadly known.

Public perception of the pandemic may also alter the frequency of searches^17^. We found that media had a strong impact on the volume of searches for taste loss and smell loss in the US, and today taste and smell loss in COVID-19 patients is rather widely known. On the other hand, in Italy the media effect on searches was slightly milder, possibly due to the advanced state of the pandemic when the news started to appear on digital media. The Italian data likely suggests genuine interest based on self-symptoms even before they became broadly known to the general public.

Interestingly, we also found a strong correlation between searches for taste loss and searches for smell loss. This is in line with recent findings that the degree of COVID-19-related anosmia and ageusia correlate closely in affected individuals^18^. Hence, both senses seem to be affected simultaneously in COVID-19 patients, a finding that may provide clues for mechanisms of action of the SARS-CoV-2 virus.

We suggest that Google Trends monitoring of taste and smell loss searches is no longer a reliable marker for COVID-19 cases, as these symptoms become common knowledge. The same limitations will likely apply to new COVID-19 symptoms that are being discovered, such as skin lesions^19^, impairment of chemesthesis (a chemosensory modality that allows the perception of burning, cooling or tingling triggered by molecules)^18^, and more^16^.

As the pandemic continues to spread around the world, and secondary waves are expected, monitoring of COVID-19 remains of great importance^20-22^ and strategies for geographic monitoring of COVID-19 hotspots are being developed^23^. Furthermore, future pandemics may, unfortunately, emerge^24^.

The phenomen described here suggests that a correlation between interest in novel symptoms of infectious disease and the number of new cases has an initial spike (the “surprise rise”) and subsequently drops off to a new baseline due to “knowledge saturation”. This lesson may be of general practical value for public health. The “surprise rise” effect has been successfully employed by several groups for outbreaks forecasting using specific Google searches. The increase in volume of searches has been shown to predict the outbreaks from several days to weeks earlier^25-27^. On the other hand, the “knowledge saturation” make the monitoring of the disease through the same tools more challenging over time.

The shortcomings of methods that rely on self-reporting should be kept in mind also in analyzing results from the various self-reporting apps^28-30^, and underscore the crucial role of independent epidemiological tools, such as sewage monitoring^31,32^ and widespread laboratory testing^23,33,34^.

## Methods

### Correlation data

The data on new COVID-19 cases per each of the 20 Italian regions were obtained from the Italian Ministry of Health website (http://www.salute.gov.it/portale/nuovocoronavirus/homeNuovoCoronavirus.jsp?lingua=english), and from the Johns Hopkins Coronavirus Resource Center for the 51 US states (https://coronavirus.jhu.edu/us-map).

The data for an individual region was normalized with respect to its total population. The number of inhabitants for each Italian region was retrieved from the last data available on the Istituto Nazionale di Statistica (National Institute for Statistics) website (http://dati.istat.it/Index.aspx?lang=en&SubSessionId=1d073136-f11a-4329-a1ed-3a3920a1ec32).

The population numbers for the US states were adopted from ^35^.

The search terms described in Table 2 were used as input in Google Trends (https://www.google.com/trends) and the data on the number of searches for each region or state were collected. Additional terms and different combinations of keywords were tested, resulting in no evident differences in the calculated correlations from the results obtained with the Table 2 terms, identified as the most popular ones. Google Trends provides the normalized number of searches according to the population living in the country’s sub-area and assigns a popularity index to the keyword searches that spans a range from 0 to 100. A value of 100 is assigned to the region in which the keyword reaches the maximum volume of searches for the dates and countries selected, with no relation to the other keywords searched in that comparison.

**Table 2:** Search terms used for searches in Italian and English on Google Trends. The first column is the reference keyword used to summarize the corresponding searches in the main text. The “+” character allows one to sum up data for multiple searches with a logical OR.

|  | Italian keywords | English keywords |
| --- | --- | --- |
| **Taste loss** | perdita gusto + perdita del gusto | taste loss + loss of taste |
| **Smell loss** | perdita olfatto + perdita dell’olfatto | smell loss + loss of smell |
| **Sight loss** | perdita vista + perdita della vista | vision loss + sight loss |
| **Hearing loss** | perdita udito + perdita dell’udito | hearing loss + loss of hearing |
| **COVID symptoms** | sintomi coronavirus + sintomi covid19 + sintomi covid + coronavirus sintomi + covid sintomi | COVID symptoms + coronavirus symptoms + COVID-19 symptoms + COVID19 symptoms + Coronavirus symptoms |

The correlation between the number of new COVID-19 cases per 1,000,000 inhabitants and the Google Trends value is Pearson’s correlation.

The collected data were used to build the graphs in Figures 1 and 2 using the RStudio software^36^.

To make sure the correlation was not dominated by an outlier in the data, a test was done on the US data by removing New York - the state with the highest number of new COVID-19 cases normalized with respect to its total population, and recalculating the correlation from the new data to find the change in correlation as insignificant.

### Media impact data

First, the popularity index of the daily queries on taste loss and smell loss from March 4th to May 19^th^ was calculated. As for the regions/states Google Trends searches, the data was automatically normalized assigning the day with the highest volume of searches the value of 100; other days were assigned values relative to that peak day.

Next, we used the Media Cloud webserver (http://www.mediacloud.org) to obtain an estimate for the number of times a certain keyword appeared on digital news on a daily basis in the period between March 4th to May 19th. By using the quoted version of the keywords as defined in Table 2 for taste loss and smell loss, the normalized numbers of appearances in digital news were obtained. The collection of media used to search our keywords were the “Italy - National” and the “U.S. Top Sources 2018” available on the Media Cloud website.

In order to superimpose the Media Cloud results to the Google Trends ones, as done in Figure 3, Media Cloud data were further normalized in the manner Google Trends are normalized: a value of 100 was assigned to the day with the highest media coverage peak of a particular search query, other days being assigned values relative to that day. The number of new cases shown in the same figure were normalized with respect to 1,000,000 inhabitants.

Figure 3 was generated using the Rstudio software ^36^.

## Data Availability

The data that support the findings of this study are openly available in "GitHub" at https://github.com/KimAsseo/Google_COVID

## Acknowledgements

We thank the Center for Interdisciplinary Data Science Research, The Hebrew University, for support, Maria Veldhuizen and Yuri Estrin for discussions, and Noam Lahav and Eitan Margulis for help in the initial stages of this project. MYN is funded by ISF grant #1129/19 and is a member of COST actions Mu.Ta.Lig (CA15135) and ERNEST (CA18133). MYN, KA, FF and JF are members the Global Consortium of Chemosensory Research, the GCCR.

## Contributions

JF initiated the research question. MYN conceived the project. MYN, KA, FF and YS suggested project direction and provided support in planning stages. KA and FF conducted data management and analysis. YS provided statistical support. KA generated the figures. FF and MYN drafted the manuscript. All authors consented to the manuscript being submitted in its final form.

## Supplementary Information

Supplementary Figure S1 repeats the calculation presented in Figure 1 for the 22-28 April week. This clearly illustrates the current lack of correlation between the number of new cases in each region (Italy) or state (US) and the popularity of Google searches on either taste or smell loss.

**Figure S1:**
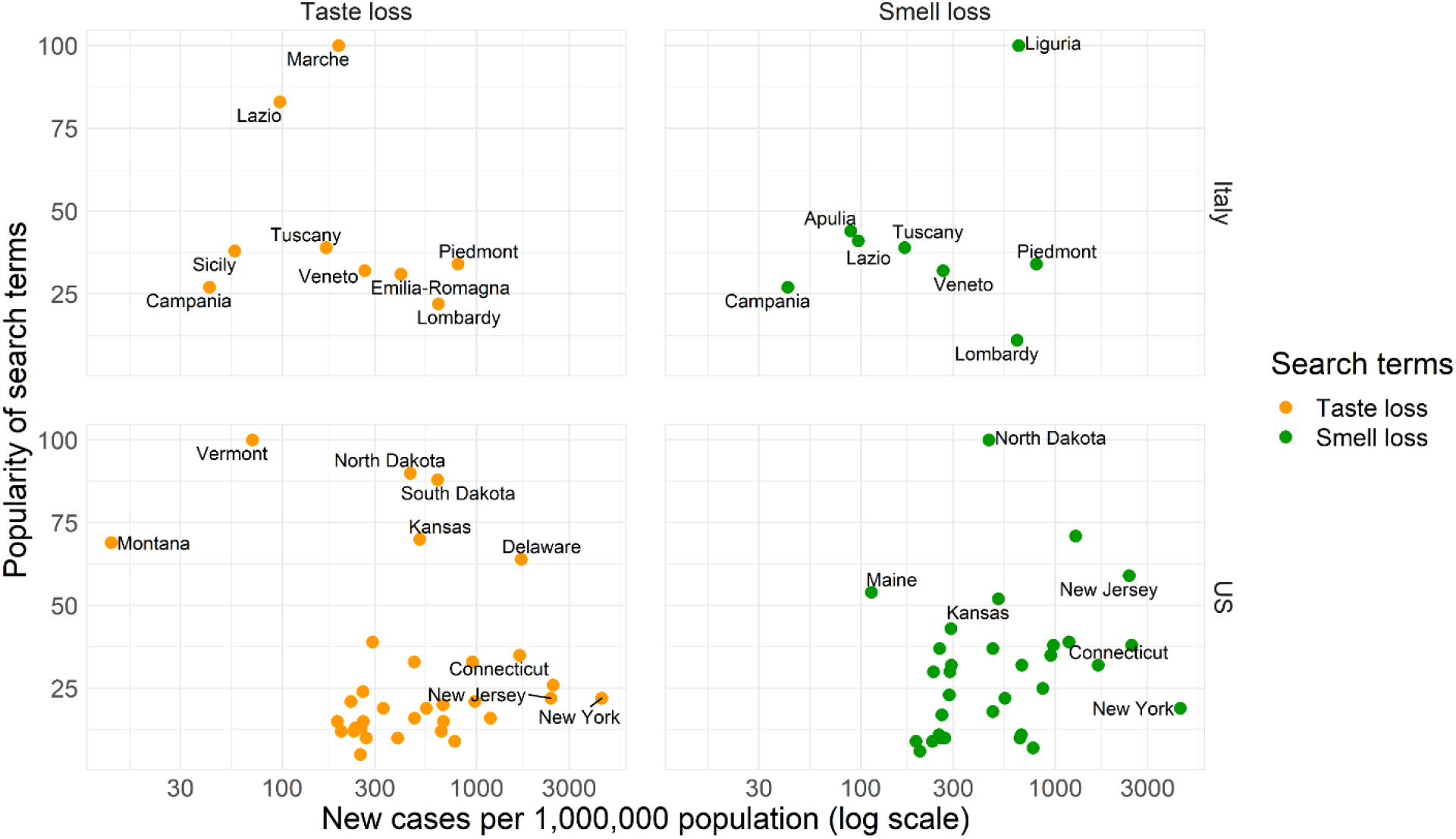
Correlation data for a low correlation week, 22-28 April 2020. The graphs describe the regions/states and include the taste loss (in orange) and smell loss (in green) search queries. Each point represents a different region/state (normalized number of new cases related to the corresponding week on the abscissa, popularity of the search terms on the ordinate axis). For both graphs, not all regions or states are shown because of the lack of popularity index for some of these geographical sub-areas.

Supplementary Figure S2 extends the period covered in Figure 3 and shows an increase in popularity of searches before the news appeared for Italy, but not for the US. It also exhibits a “surprise rise” in both countries with media coverage and new cases rising, followed by a “knowledge saturation”, where the baseline of searches is somewhat higher than before, but overall low and not reflective of the number of new cases in the respective country.

**Figure S2:**
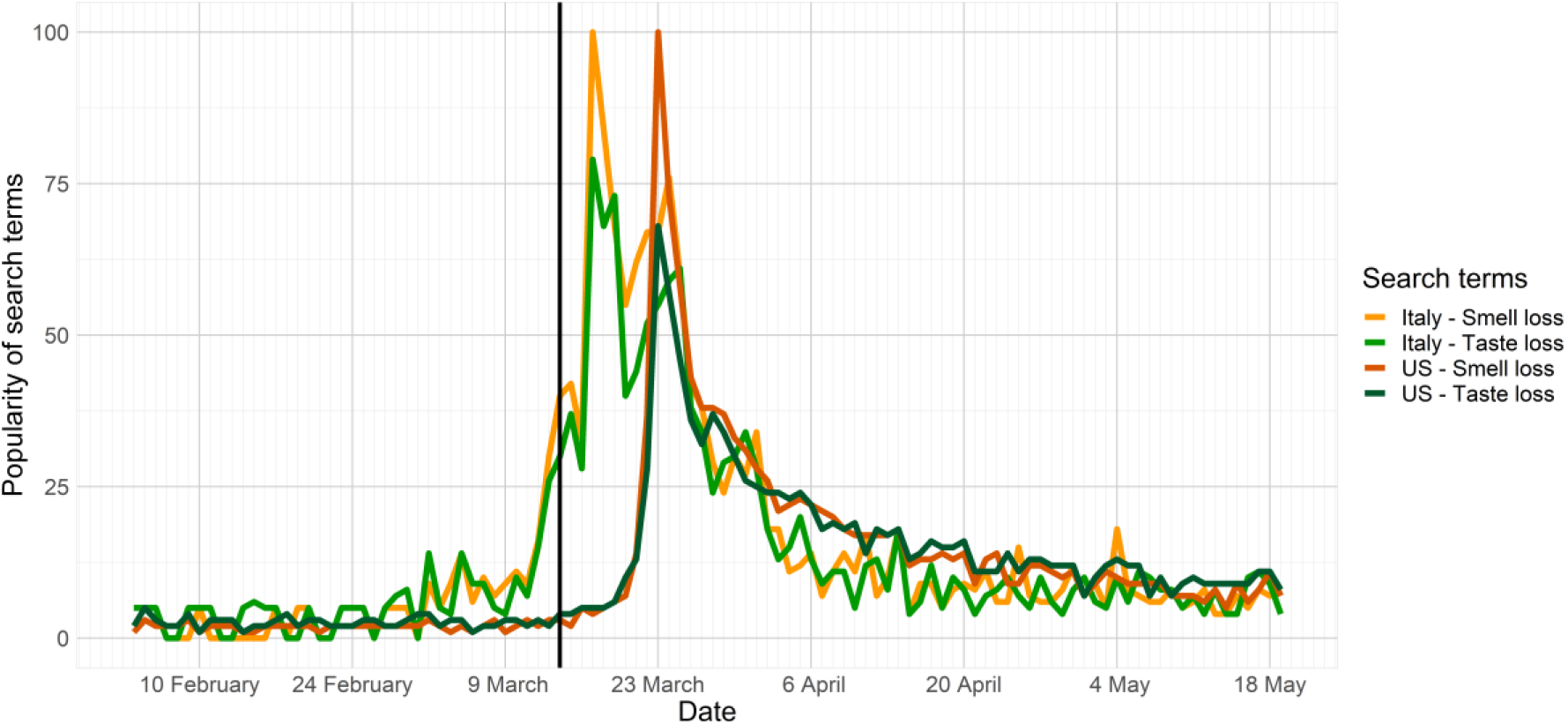
Popularity of smell loss and taste loss searches for Italy and the US from February 4^th^ to May 19th. The former date lies one month before the point in time from which the records have been analyzed in the present study. The date the phenomenon was reported in the media for the first time, on March 14^th^, 2020, is shown as a black vertical line.

